# Computational Strategies for Depression Detection and Treatment: The Role of Behavioral Activation and Neurobiological Insights – A Systematic Review

**DOI:** 10.1101/2025.03.27.25324813

**Authors:** Paula Tallón Fuentes, Amaia Méndez Zorrilla, Markus Reichert, Begoña García-Zapirain Soto

## Abstract

**Objective:** Depression is a multifaceted disorder with neurobiological, behavioral, and environmental components. This review aims to explore how artificial intelligence (AI) and computational methods are advancing the understanding and treatment of depression, focusing on neurobiological mechanisms, early detection, and behavioral activation (BA) interventions.

**Methods:** A comprehensive literature review was conducted searching PubMed, Scopus, ACM, and Web of Science databases. From 77654 articles identified, 48 studies were selected based on relevance and methodological rigor. These include meta-analyses, randomized controlled trials, or observational studies, focusing on the integration of AI and computational tools in depression research and treatment.

**Results:** Advances in AI-driven neuroimaging and machine learning have enhanced the identification of neurobiological changes associated with depression, such as hippocampal atrophy, prefrontal cortex dysfunction, and HPA axis dysregulation. AI models have also facilitated early detection of subtle biomarkers linked to neuroinflammation and reduced BDNF levels. Furthermore, AI-powered digital platforms have optimized BA interventions, personalized treatment and improving access through virtual coaching and mobile applications. AI-enhanced interventions incorporating physical activity monitoring have shown neuroprotective effects, promoting neurogenesis, reducing inflammation, and increasing BDNF levels.

**Conclusion:** The integration of AI and computational approaches into traditional depression therapies holds significant promise. AI-driven tools, when combined with BA interventions, provide scalable, personalized solutions, particularly for individuals with limited access to conventional treatments. The future of depression care can strongly profit from the convergence of AI, neurobiology, and behavioral science, which will enhance diagnostic accuracy, treatment effectiveness, and accessibility.

## 1. INTRODUCTION

Depression is one of the leading mental health conditions worldwide, affecting around 246 million people. It presents itself in various forms, from major depressive disorder (MDD) to seasonal affective disorder (SAD) and other mood-related disorders. It is characterized by persistent feelings of sadness, loss of interest in daily activities, and physical symptoms such as changes in appetite, sleep disturbances, and cognitive impairments Different studies have explored various factors that contribute to the onset and persistence of depression, including physical inactivity, aging, brain health, and behavioral factors (1)Behavioral activation (BA) is an effective therapeutic approach for treating depression. BA focuses on helping individuals engage in meaningful activities that bring a sense of accomplishment or pleasure. It has been shown to be particularly effective in treating depression by addressing the inactivity and withdrawal that often accompany depressive episodes (2). BA can be enhanced when combined with physical activity, as exercise not only helps to improve mood but also provides an opportunity for individuals to engage in structured activities that promote well-being (1).

PA is defined as “any bodily movement produced by skeletal muscles that requires energy expenditure”. The relationship between depression and PA is reciprocal—depression reduces PA levels, and low PA levels exacerbate depression. By enhancing neuroplasticity, improving neurochemical balance, and reducing stress and inflammation in the brain, PA has become an promising part of mental health treatment plans, alongside traditional therapeutic interventions such as medication and psychotherapy (1).

One of the key risk factors for depression is frailty. As individuals age, they may experience declines in physical function, mobility, and social engagement, which can lead to feelings of helplessness, loneliness, and depression. The relationship between frailty and depression is complex and bidirectional; frailty can exacerbate depressive symptoms, while depression can accelerate the onset of frailty through decreased physical activity and social withdrawal (3). Interventions that address both frailty and depression, such as physical rehabilitation and social support, have shown promise in improving the quality of life for elderly individuals (3).

Neuroimaging studies have revealed that depressive disorders are often associated with structural and functional changes in the brain, particularly in areas related to mood regulation, such as the prefrontal cortex and hippocampus. These changes may contribute to the emotional dysregulation and cognitive impairments commonly observed in depression (4). Research suggests that interventions such as exercise, cognitive behavioral therapy (CBT), and pharmacotherapy may help reverse some of these brain changes, improving both brain function and mental health (2).

Advancements in computational tools and data analysis techniques have paved the way for more accurate and objective measures of depression. Brain-computer interfaces (BCIs), machine learning algorithms, and neuroimaging have been increasingly applied to improve diagnosis and understanding of depression. For instance, recent studies have shown how brain-computer interfaces can use electroencephalogram (EEG) signals to detect depression patterns with enhanced accuracy, offering an alternative to traditional self-report measures (5). Others have utilized deep learning models and neuroimaging to improve diagnostic precision and understand the neural correlates of depression (6,7). These technologies provide insights into the neural mechanisms underlying depressive states, enhancing the ability to identify and treat depression more effectively.

An increasing body of research has highlighted the complex interplay between physical activity, frailty, brain health, and depression. Studies have shown that physical activity can help mitigate the effects of frailty, improve brain health, and enhance the efficacy of behavioral activation strategies in treating depression (1,8). The integration of these factors into a unified approach to mental health, along with the use of advanced data analysis tools, has the potential to improve therapeutic outcomes for individuals suffering from depression (2). The application of computational methods such as multimodal data fusion, deep learning, and network analysis is shaping the future of depression research and treatment, making the diagnosis and management of depression more accurate and efficient This review aims to explore the interconnectedness of the previous mentioned concepts, and examines how these factors can be leveraged to create more effective interventions for depression. By synthesizing the findings of numerous studies, we aim to provide a comprehensive understanding of how an integrated approach can improve the management and treatment of depression, ultimately leading to better outcomes for individuals affected by this debilitating condition.

## 2. MATERIALS AND METHODS

**Table 1.** Research questions.

| Research question | Purpose |
| --- | --- |
| **Q1: What are the device-based metrics used to quantify the behavioral activation and its influence in reduction of depressive symptoms?** | To explore the effectiveness of behavioral activation (including activity scheduling and task initiation) in reducing depressive symptoms in adults with major depressive disorder, and its potential as a therapeutic approach. |
| **Q2: What are the parameters obtained after applying signal processing techniques to neurobiological and structural brain images to determine which are associated with depression?** | To examine which specific brain structures and functions (e.g., prefrontal cortex, hippocampus) are altered in individuals with depression, and how these changes correlate with different phenotypic responses to depression treatments, including physical activity and behavioral activation. |
| **Q3: How do frailty and other individual and environmental factors (e.g., age, comorbidities) influence the effectiveness of those interventions? Which is the correlation between frailty and depression?** | To determine the role of frailty and other health conditions in the reduction of the impact of the treatment interventions on depression. |
| **Q4: What are the current computational approaches, including artificial intelligence, in the detection and monitoring of depression?** | To explore and clarify the current ways, in computational terms, of obtaining, processing and analysing data from patients and to determine if it's an interesting approach in order to accelerate and improve the current treatments. |

### 2.1 Data Collection

In conducting this review, we adhered to the Preferred Reporting Items for Systematic Reviews and Meta-Analyses (PRISMA) 2020 guidelines to ensure a comprehensive and transparent approach to data collection. Our search initially yielded a total of 77,654 articles sourced from a variety of databases. These articles focused on a range of topics, including the intersection of physical activity, depression, frailty, and mental health interventions in both aging and adult populations, with a specific emphasis on neuroimaging, behavioral activation therapies, and the impact of exercise on mental well-being.

Through careful selection, we included studies that examined physical activity interventions, the role of depression in frailty, and emerging machine learning models for depression diagnosis, ensuring a robust representation of the existing literature on these interconnected domains.

#### 2.1.1 Searched databases

The following databases were systematically searched for relevant studies: Web of Science, Scopus, ACM and PubMed. These databases were selected due to their comprehensive coverage of peer-reviewed scientific literature in the fields of psychology, neuroscience and physical health, and they are commonly used for research in the domain of depression and related conditions.

#### 2.1.2 Searched term**s**

The terms used in the searches included combinations of keywords related to depression, physical activity, frailty, informatics, computer-based tools, brain phenotypes, and behavioral activation. The specific search chains used across the databases were:

- "depression AND brain phenotype"
- "depression AND behavioral activation AND physical activity"
- "depression AND physical activity"
- "depression AND frailty"
- "depression AND frailty AND physical activity"
- "depression AND major depressive disorder"
- "depression AND brain phenotype"
- "behavioral activation AND major depressive disorder"
- “behavioral activation AND computational AND depression”
- “behavioral activation AND neural networks AND depression”
- “depression AND neural networks AND brain”
- “neurological AND computational AND depression”
- “depression AND neural networks AND brain phenotype”

#### 2.1.3 Inclusion criteria

This review focused on studies that investigated the integration of artificial intelligence (AI) and computational methods in understanding and treating depression. Eligible studies were required to meet the following criteria: (1) they must have involved AI-driven neuroimaging, machine learning, or digital interventions targeting depression, (2) they should have addressed neurobiological mechanisms, such as brain structural changes or biomarkers linked to depression, (3) studies must have used robust methodologies, including meta-analyses, randomized controlled trials, or observational studies, (4) the studies should have focused on behavioral activation interventions enhanced by AI or AI-supported platforms for early detection and personalized treatment, (5) studies from peer-reviewed journals were considered for inclusion. Studies with a primary focus on non-depression disorders, systematic reviews, opinion articles or ones lacking methodological rigor were excluded.

#### 2.1.4 Quality assessment and bias

To further ensure the rigor and reliability of the included studies, we applied the Cochrane Risk of Bias (RoB 2) tool for randomized controlled trials (RCTs), the AMSTAR 2 (A Measurement Tool to Assess Systematic Reviews) for systematic reviews and meta-analyses. The RoB 2 tool evaluates studies based on five key domains: (1) bias arising from the randomization process, (2) bias due to deviations from the intended interventions, (3) bias due to missing outcome data, (4) bias in measurement of the outcome, and (5) bias in selection of the reported result. Each domain was rated as low risk, some concerns, or high risk of bias, and an overall risk rating was assigned accordingly.

For systematic reviews and meta-analyses, AMSTAR 2 was employed to assess methodological quality based on 16 criteria, including protocol registration, adequacy of the literature search, risk of bias assessment in included studies, and appropriateness of statistical methods.

In addition, observational studies were evaluated using the Newcastle-Ottawa Scale (NOS), which assesses study quality based on selection, comparability, and outcome measures.

For studies involving machine learning (ML) methodologies, we applied the Minimum Information for Medical AI Reporting (MINIMAR) framework, as recommended by Nature Medicine. Traditional tools like Cochrane RoB 2 and AMSTAR 2 do not fully capture the reproducibility challenges of AI models.

Therefore, this tool was used to assess different aspects such as dataset size, external validation, risk of overfitting and transparency in reporting model performance.

The risk of bias findings was incorporated into the summary tables of included studies, with an additional column indicating the risk of bias level (low, moderate, or high) for each study. This approach ensures a more transparent evaluation of study reliability and allows for a more nuanced interpretation of the findings in the results and discussion sections.

All results from our search strategy were initially screened for relevance by the first author (PT) based on title and abstract. The papers that were found to be relevant underwent full evaluation for eligibility by PT and BG independently, and the results from both investigators were compared. In cases where one of the investigators was uncertain about including a paper, two additional authors (MR and AM) reviewed the full article. All reviewers then re-evaluated the article to reach a consensus on its inclusion.

Overall, while there was a fair level of quality in the studies, potential sources of bias, including publication bias and the small sample sizes in some studies, were considered in our analysis. The limitations stemming from the quality of the included studies and missing data were further addressed in the discussion section, where we provided recommendations for future research. The systematic review was conducted following the principles and guidelines of the PRISMA model.

To facilitate the selection of the most relevant papers, a metric table was made, taking in consideration qualities such as the design of the study, the impact and others such as the proper quality in terms of content (Table 2).

**Table 2.** Paper quality metrics.

| Metric type | Item | Description | Values | Weight |
| --- | --- | --- | --- | --- |
| Study design | 1 | Originality of the study design (experimental, observational, systematic review, meta-analysis). | Observational (1), Experimental (2), Review (0.5) | 2 |
|  | 2 | Sample size adequacy based on study objectives. | Small (<50, 0.5), Medium (50-200, 1), Large (>200, 2) | 1 |
| Content Quality | 3 | Clear description of methodology (e.g., data collection, intervention, statistical analysis). | Yes (1), No (0) | 2 |
|  | 4 | Relevance to research questions (e.g., focus on physical activity, depression, frailty). | Low (0.5), Moderate (1), High (2) | 2 |
| Findings & Implications | 5 | Includes findings on neurobiological, behavioral, or social factors influencing depression. | Yes (1), No (0) | 2 |
|  | 6 | Addresses frailty or brain-related phenotypes as part of the analysis. | Yes (1), No (0) | 1 |
|  | 7 | Contribution to understanding of treatment strategies for depression. | Low (0.5), Moderate (1), High (2) | 2 |
| Transparency | 8 | Availability of data or code for replication. | Yes (1), No (0) | 0.5 |

| **Metric type** | **Item** | **Description** | **Values** | **Weight** |
| --- | --- | --- | --- | --- |
|  | 9 | Provides clear discussion of limitations and biases. | Yes (1), No (0) | 1 |
| Citation Impact | 10 | Cited by other studies beyond self-references. | Yes (1), No (0) | 0.5 |

## 3. RESULTS

After screening and selection, 272 items were selected and included in the Zotero database (Figure 1.). Subsequent to a detailed content review, following the indicated criteria, 116 articles were selected for further analysis based on their relevance. To ensure the highest quality and relevance to the study’s focus, we thereafter applied a metric-based approach to prioritize the articles, ultimately selecting 54 articles for inclusion. The number of articles selected by year is collected in the frequency diagram (Figure 2).

**Figure 1.**
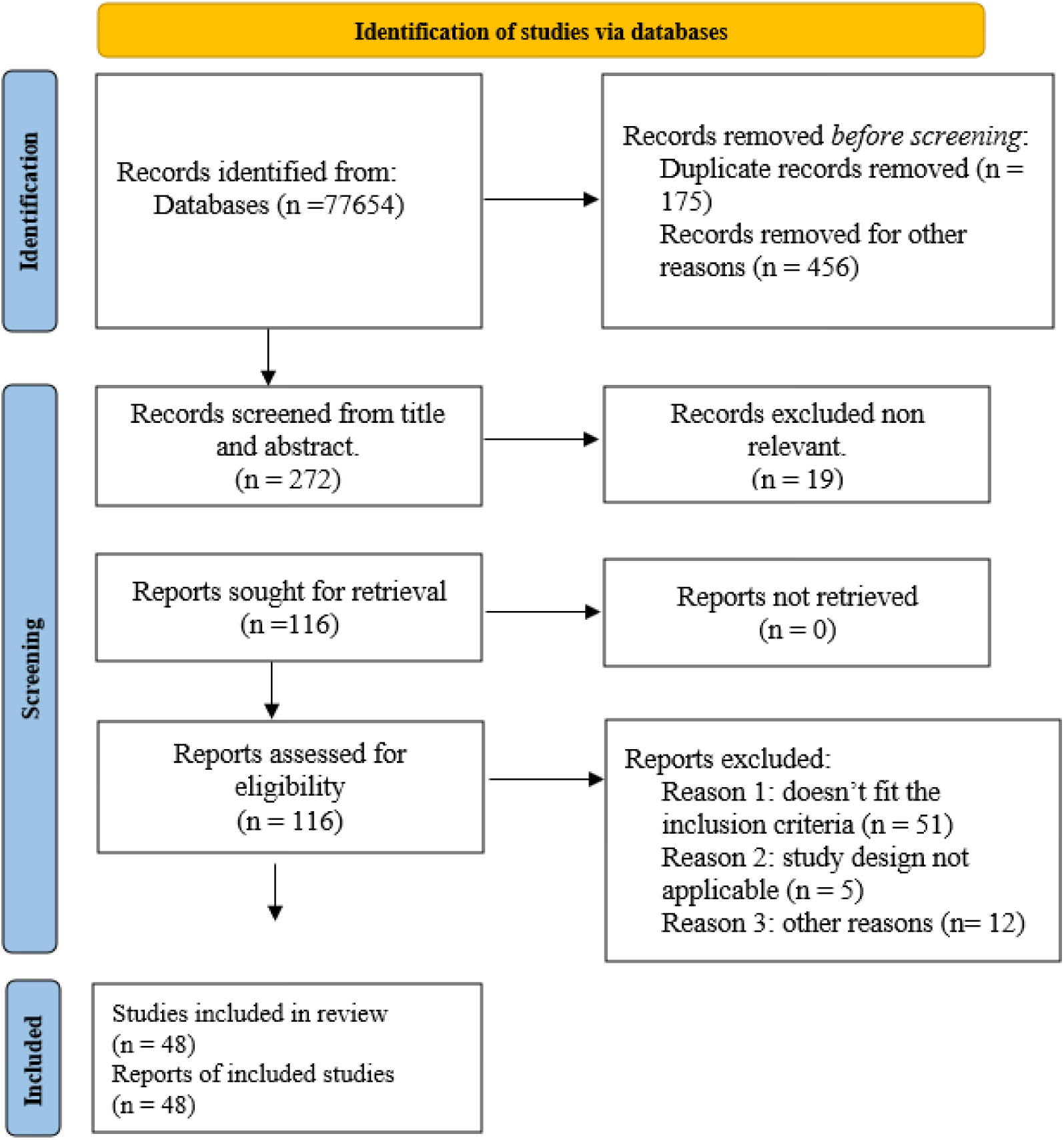
PRISMA 2020 flow diagram for included searched articles from databases.

**Figure 2.**
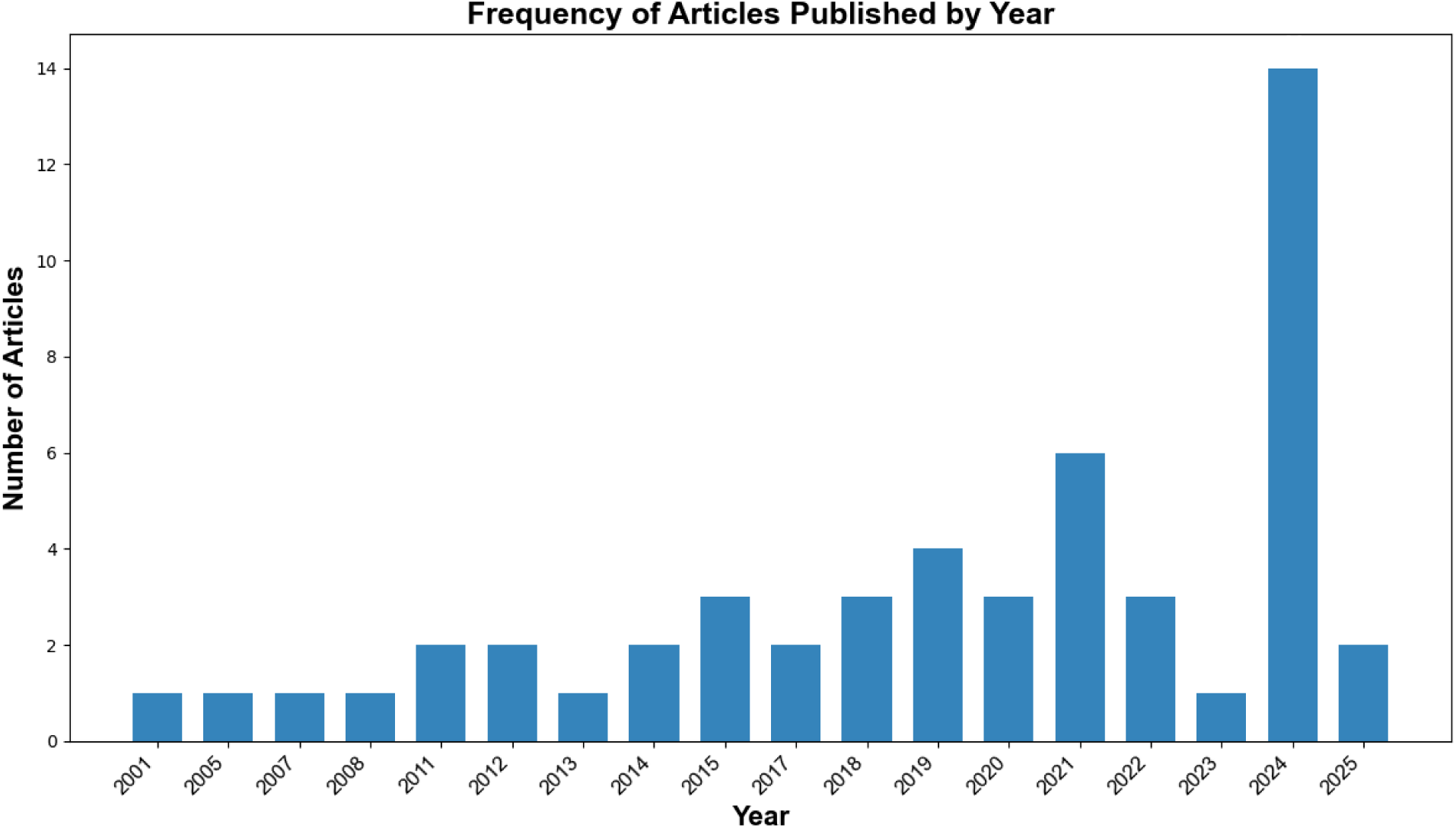
Number of articles selected and year of publication.

### 3.1 Description of Included Studies

The tables below (Table 3) summarize the main characteristics of the studies that were included in this review. The table includes author(s), year of publication, study design, sample size, population, methodology or intervention, key outcomes, and conclusions. Additionally, the research question addressed by each study and the risk of bias (low, moderate, or high) are presented.

**Table 3.**
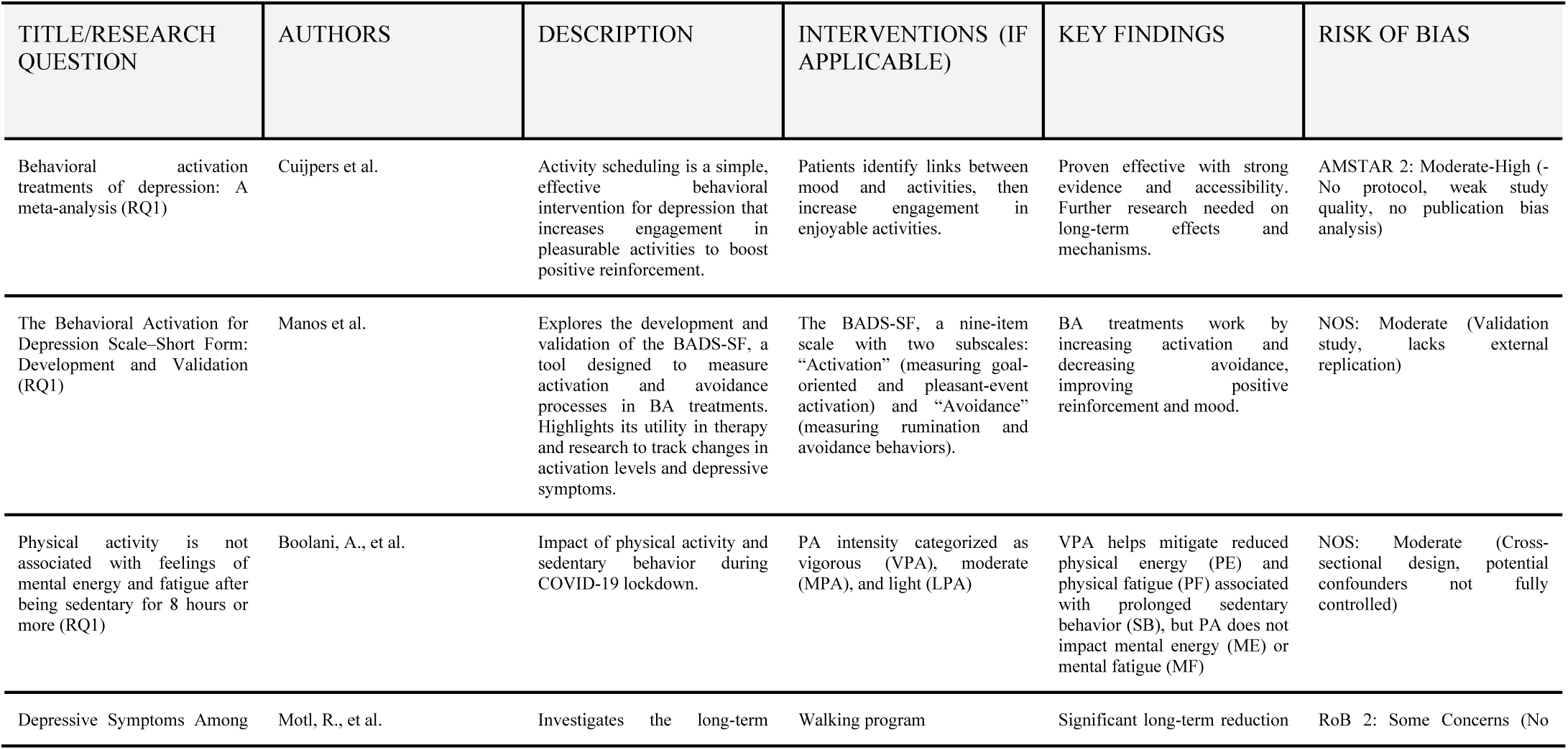
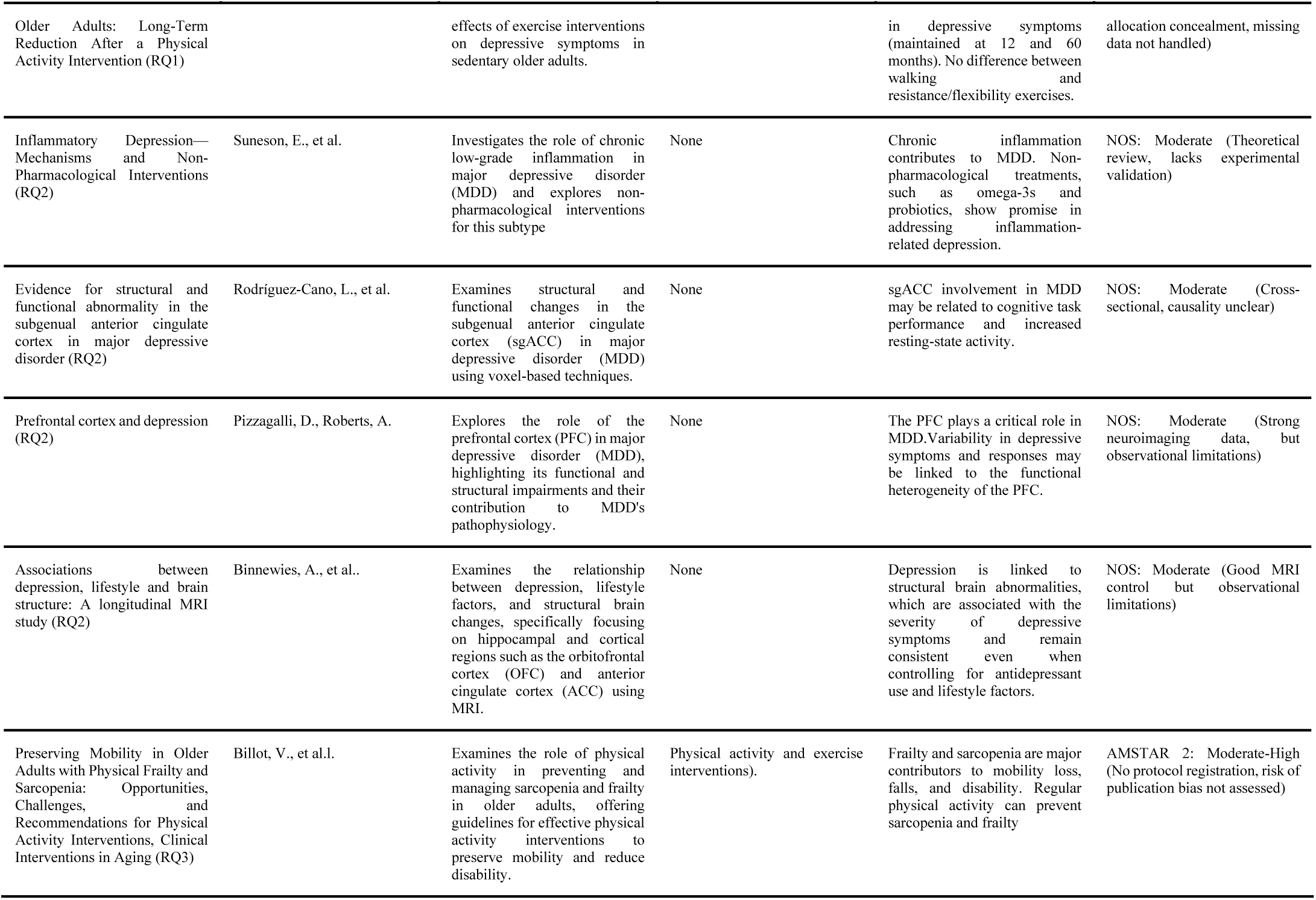
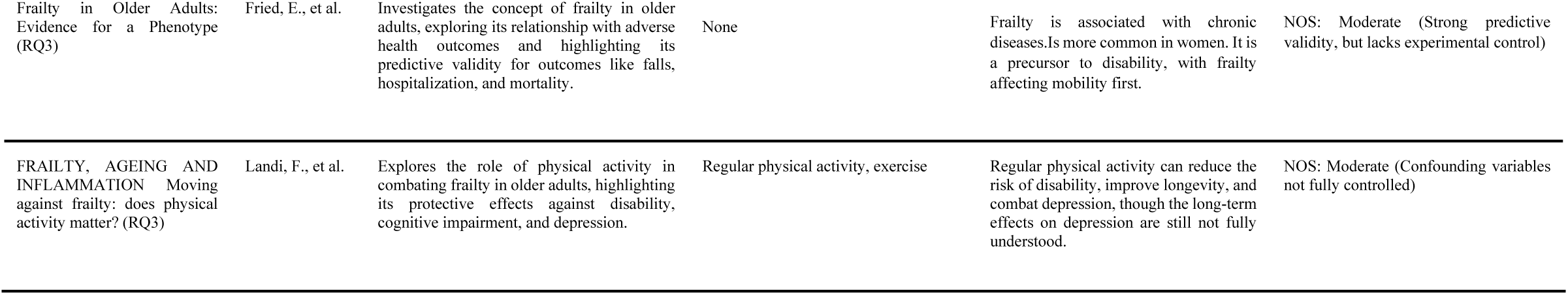
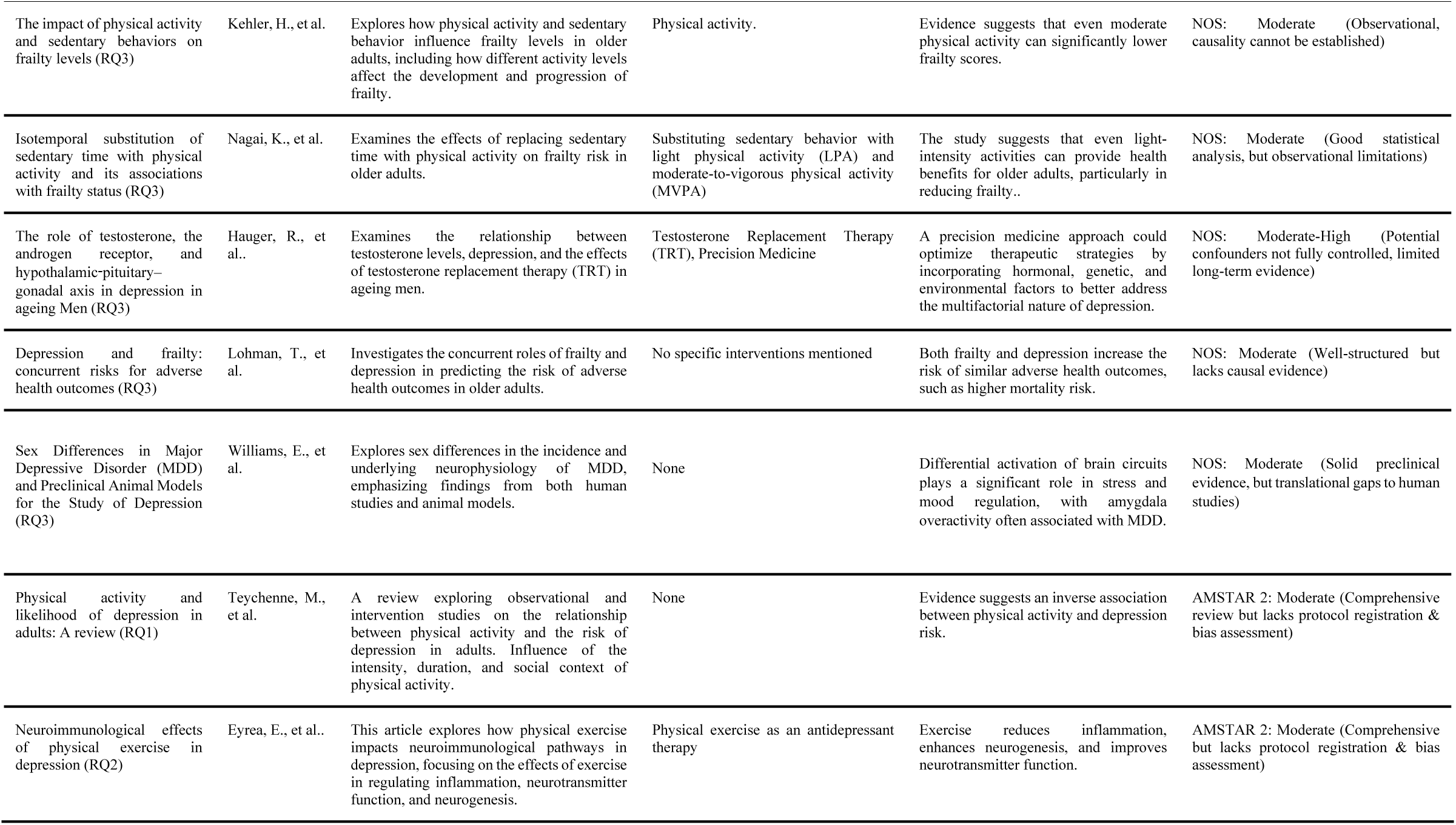
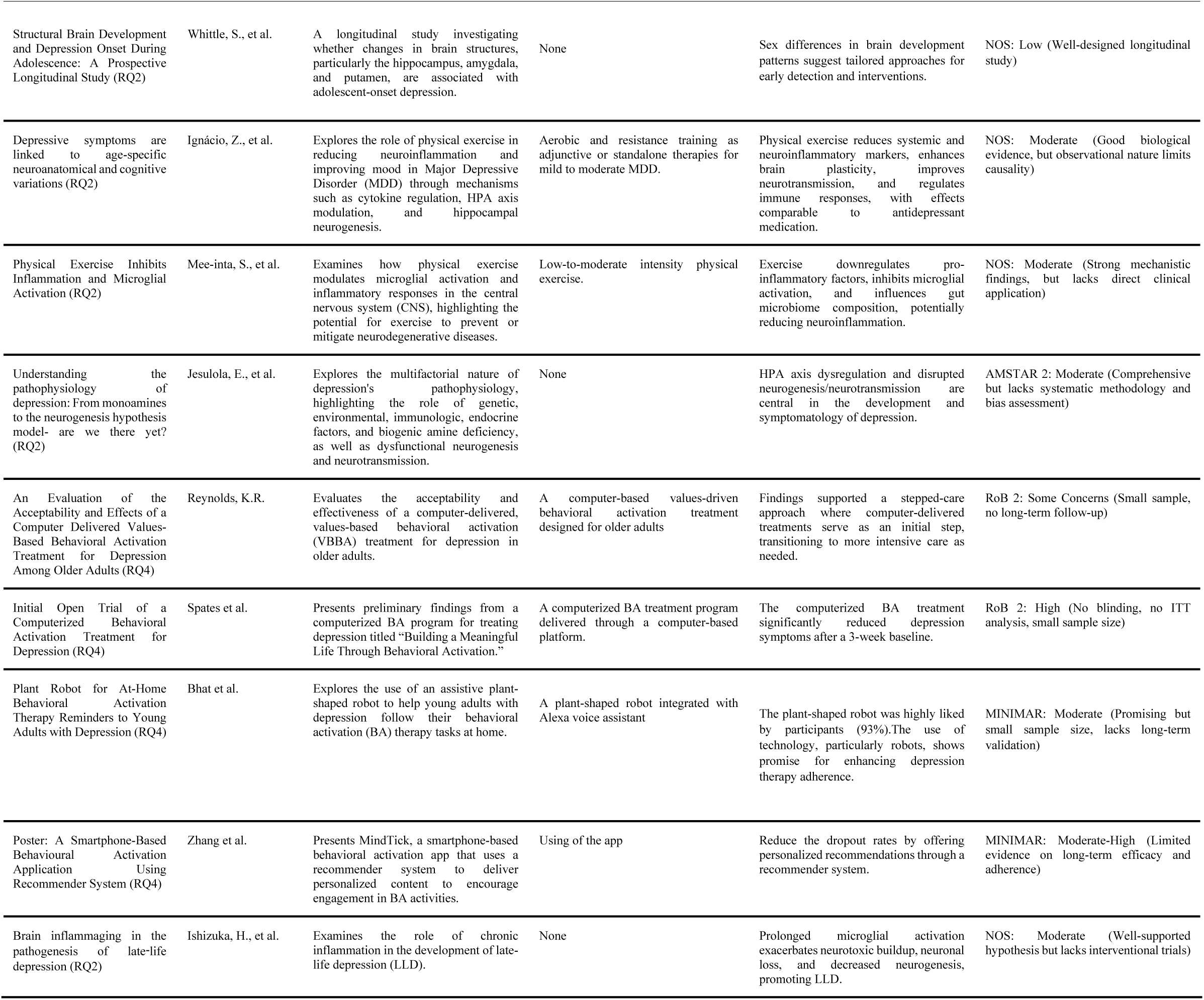
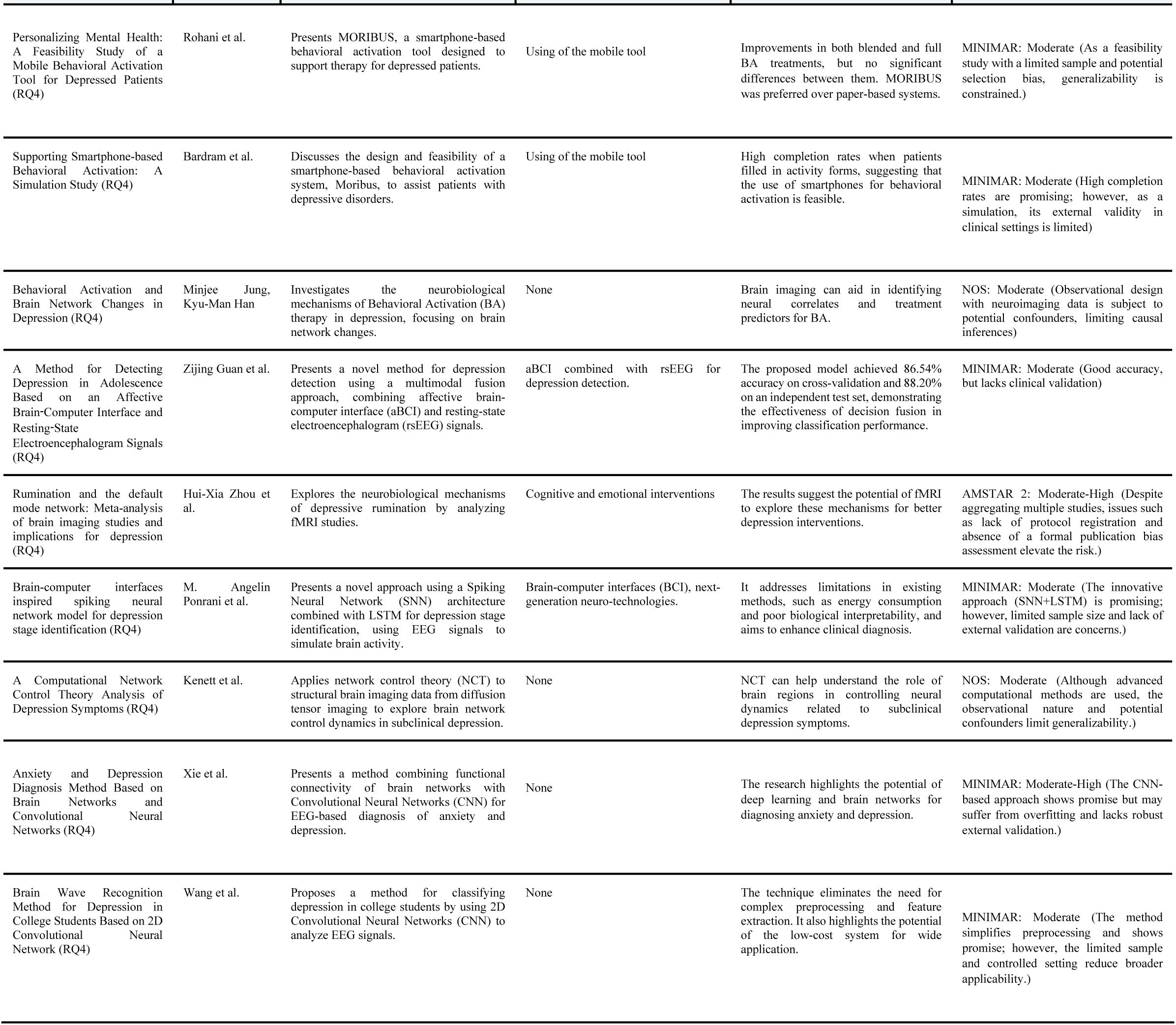
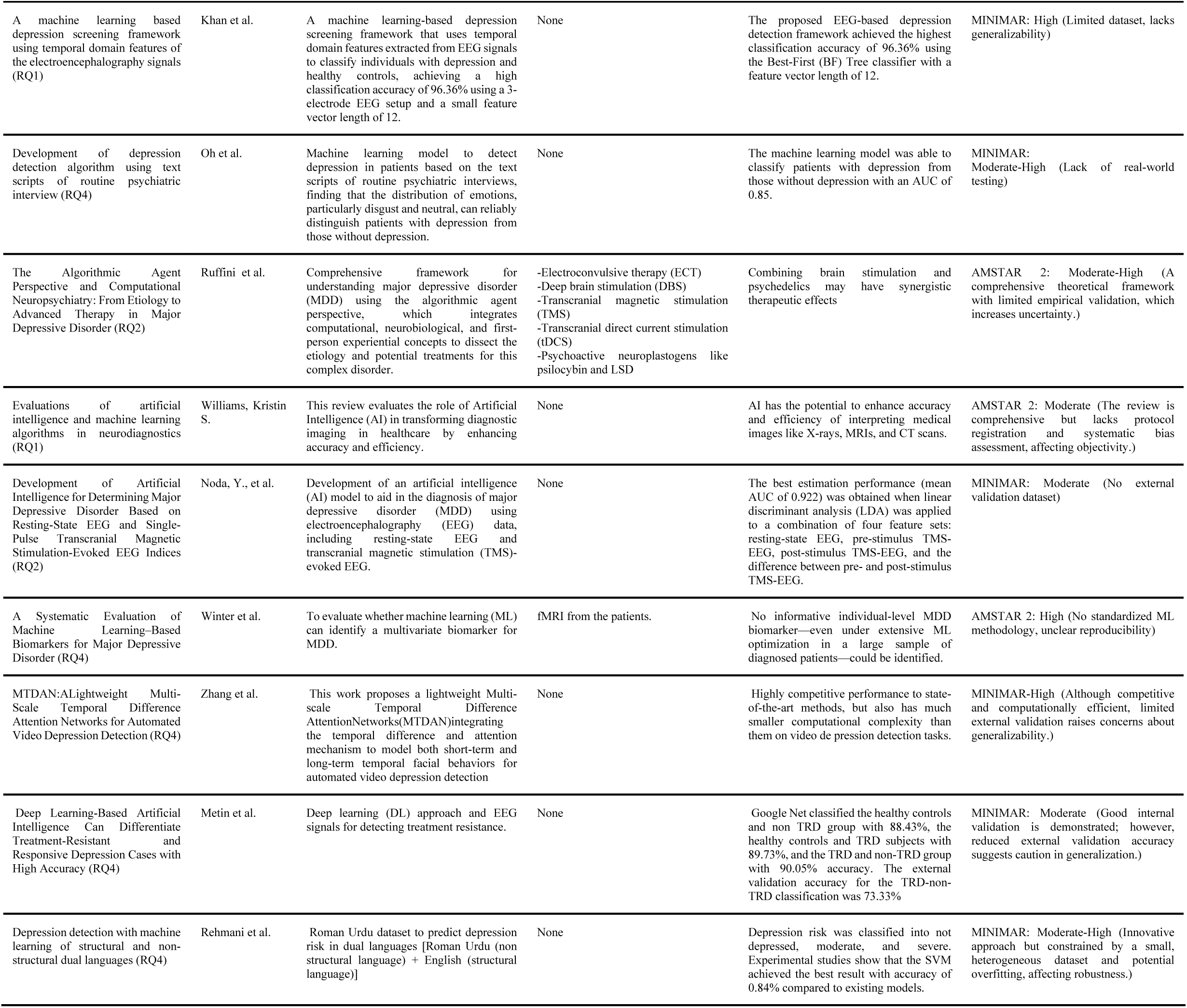
Summary table with the main characteristics of the studies and the research question associated.

### 3.2 Effectiveness of Behavioral Activation (BA)

Behavioral Activation (BA) has emerged as a structured psychotherapeutic approach with proven efficacy in treating depression by focusing on increasing engagement in rewarding activities and reducing behaviors that reinforce depressive symptoms. Studies have consistently demonstrated the therapeutic benefits of BA in addressing the core mechanisms of depression, particularly through activity scheduling and structured engagement. This section synthesizes the evidence on the effectiveness of BA, with an emphasis on its interplay with physical activity and its application across different populations.

#### 3.2.2 BA and Physical Activity: A Synergistic Approach

BA’s mechanism of change involves increasing behavioral activation while reducing avoidance behaviors (9) thereby enhancing contact with positive reinforcement. Over time, this repeated cycle strengthens positive mood and mitigates depressive symptoms. Despite its proven efficacy, research continues to explore the nuances of BA, including its long-term outcomes and optimal delivery methods.

Physical activity (PA) serves as a natural complement to BA, providing structured opportunities for positive reinforcement while also addressing physiological and neurobiological contributors to depression. The integration of BA with PA interventions has been particularly effective in older populations, where the interplay between depression, frailty, and sedentary behavior is pronounced. Regular physical activity, including light incidental movements, helps mitigate symptoms of depression in older adults, while sedentary behavior exacerbates these symptoms by fostering physical and mental fatigue (10). These findings underscore the importance of incorporating BA strategies into physical activity promotion, particularly for populations at risk of chronic inactivity or social isolation.

Experimental evidence supports PA as a behavioral intervention with antidepressant effects comparable to cognitive-behavioral therapy or pharmacological treatments(11). However, challenges remain, such as determining the optimal intensity and duration of PA to maximize outcomes (12). Additionally, addressing barriers to activity engagement, such as physical frailty or environmental constraints, is critical for ensuring the long-term success of BA interventions.

#### 3.2.3 Integration of Computer-Based Programs in Behavioral Activation

Recent advancements have made use of technological innovations, particularly computer-based programs and mobile applications, to expand the reach and effectiveness of BA. This section reviews several studies that explore the integration of computer-delivered and mobile-based interventions in the context of BA, highlighting their potential benefits and challenges.

One key example is the "Building a Meaningful Life Through Behavioral Activation" (BAML) program (13). This computerized intervention was tested in a preliminary open trial, which showed significant reductions in depression symptoms after just a three-week baseline period. Participants in the trial also reported substantial improvements in their overall quality of life, with these positive effects being maintained at a 6-month follow-up. These findings demonstrate the potential of computer-delivered BA interventions in reaching individuals who may not have access to traditional face-to-face therapy.

##### 3.2.3.1 Assistive Technologies in Behavioral Activation

Alongside computerized programs, mobile applications have become an important avenue for delivering BA interventions. A notable example of a mobile app designed specifically for BA is the MindTick app (6). This app uses a recommender system to suggest personalized activities based on users’ previous interactions and preferences. MindTick encourages users to engage in daily activities and provides tools for self-monitoring, action planning, and mood tracking. Early results suggest that this personalized approach may reduce dropout rates compared to more generic applications and can enhance user engagement, which is often a challenge in digital interventions for mental health.

In addition to traditional digital tools, assistive technologies such as robots have been integrated into BA interventions to further enhance engagement and adherence. One example is a plant-shaped robot (14), which was designed to provide reminders for completing BA tasks. This innovative design was specifically targeted at young adults, a population that is particularly prone to disengagement with mental health treatments. The plant-shaped robot proved to be a popular choice, with 93% of participants expressing enjoyment and satisfaction with the design. The results from this prototype suggest that incorporating personalized, non-traditional designs can significantly improve adherence to BA tasks.

Another promising development in the integration of technology into BA therapy is the use of personalized interventions. The MORIBUS tool (15), uses smartphones to track patients’ activities and emotions, providing them with visual feedback and insights into their behavior patterns. This tool aims to personalize BA by tailoring interventions based on real-time data, allowing patients to better understand their emotional and behavioral trends. Research on this tool suggests that mobile versions of BA are more engaging and efficient than paper-based approaches, with users showing a preference for smartphone-based tracking systems over traditional methods.

##### 3.2.3.2 Neurobiological Insights into Behavioral Activation and Technology

Although behavioral activation interventions are primarily psychological, there is growing interest in understanding the neurobiological mechanisms underlying these therapies, and therefore, there is a need for neuroimaging studies to explore how BA influences brain activity in individuals with depression (16). While cognitive behavioral therapy (CBT) has been studied extensively in this context, research into the neurobiological effects of BA remains sparse. Investigating these mechanisms could provide valuable insights into how technology-based BA interventions might influence brain networks, offering a more comprehensive understanding of the treatment’s effectiveness and potential biomarkers for identifying patients who would benefit most from BA.

### 3.3 Neurobiological and Structural Brain Changes

The neurobiological mechanisms underlying depression are multifaceted, involving significant alterations in brain structure, neurochemical systems, and neuroinflammatory processes. These changes have been implicated in both the onset and progression of depression, with notable differences observed across various stages of life, particularly between late-life depression (LLD) and early-life depression (ELD). Structural and functional brain changes, as well as disruptions in neurochemical and neuroinflammatory pathways, are key features that contribute to the symptoms of depression, including cognitive dysfunction and emotional dysregulation(17,18).

#### 3.3.1 Structural Brain Changes

Neuroimaging studies using structural magnetic resonance imaging (sMRI) have revealed consistent alterations in the size and volume of specific brain regions involved in mood regulation, stress responses, and cognitive function. These include the hippocampus, prefrontal cortex (PFC), amygdala, and anterior cingulate cortex (ACC), which are crucial in emotional processing and cognitive functions. Studies show that the hippocampus, in particular, is a region that demonstrates significant volume reductions in individuals with depression, especially in those suffering from chronic or late-onset depression (19,20)These reductions in hippocampal volume are believed to contribute to the emotional dysregulation and cognitive impairments characteristic of depression (21).

Further changes are observed in the prefrontal cortex (PFC), particularly in the ventromedial and dorsolateral areas. These regions play a critical role in executive functions, decision-making, and regulating emotional responses. Structural changes in these regions are frequently associated with depressive symptoms, including cognitive dysfunction and difficulty in regulating mood (22). Additionally, reductions in the volume of the anterior cingulate cortex (ACC), particularly the subgenual ACC, are consistently reported in major depressive disorder (MDD), with decreased ACC activity correlating with heightened emotional reactivity and impaired regulation (17,23).

#### 3.3.2 Neurochemical and Inflammatory Pathways

In addition to structural changes, neurochemical alterations, particularly within the monoaminergic systems—serotonin, dopamine, and norepinephrine—are crucial in the pathophysiology of depression. The disruption of these neurotransmitters, in conjunction with altered neuroinflammatory processes, underlies many of the clinical manifestations of depression. Proinflammatory cytokines such as IL-6 and TNF-alpha, along with neurotrophins like BDNF, have been shown to be elevated in depressed individuals, indicating a critical role for neuroinflammation in the development and persistence of depressive symptoms (11)

There is growing evidence that the neuroinflammatory response, particularly in the hippocampus and prefrontal cortex, contributes to neuronal dysfunction and reduces neuroplasticity in depression (24). This inflammatory process is also associated with impaired neurogenesis, particularly in the hippocampus, where neurogenesis is known to be crucial for learning, memory, and emotional regulation (25,26)The interaction between the hypothalamic-pituitary-adrenal (HPA) axis and inflammatory pathways further exacerbates depression’s neurobiological underpinnings. Chronic stress and hyperactivity of the HPA axis lead to elevated cortisol levels, which, over time, contribute to hippocampal atrophy and functional impairments (11,24) Moreover, microglial activation, which is commonly seen in the brains of individuals with depression, further amplifies the inflammatory response, leading to a self-perpetuating cycle of neuroinflammation and neuronal damage (25,27)

#### 3.3.3 Neuroprotective Effects of Exercise

Physical exercise has emerged as a potential intervention to counteract the neurobiological changes associated with depression. Regular physical activity has been shown to reduce inflammation and promote neurogenesis, particularly in the hippocampus, offering neuroprotective benefits for individuals with depression. Exercise has been linked to reductions in proinflammatory cytokines and increased levels of brain-derived neurotrophic factor (BDNF), which supports neuroplasticity and cognitive function (20,26)Furthermore, studies indicate that exercise may enhance the functioning of the prefrontal cortex, facilitating improved emotional regulation and cognitive control in individuals suffering from depression (17,19).

Neurobiological and structural brain changes in depression involve complex interactions between alterations in brain volume, neurotransmitter systems, and neuroinflammation. These changes not only provide insights into the underlying mechanisms of depression but also suggest potential therapeutic targets. Reducing inflammation and promoting neurogenesis through interventions such as physical exercise offer promising avenues for treating the neurobiological deficits observed in depression. Understanding these pathways is essential for the development of more effective, individualized treatments for depression (26,28).

#### 3.3.4 Advances in Data Analysis and Computational Tools in Neurological Research

Recent developments in the application of computational tools and data analysis techniques (Figure 3., Figure 4.) have significantly advanced depression research, particularly in the field of affective neuroscience and mental health diagnostics. Several studies have highlighted the promising role of brain-computer interfaces (BCIs) and advanced machine learning techniques in detecting and understanding depression.

**Figure 3.**
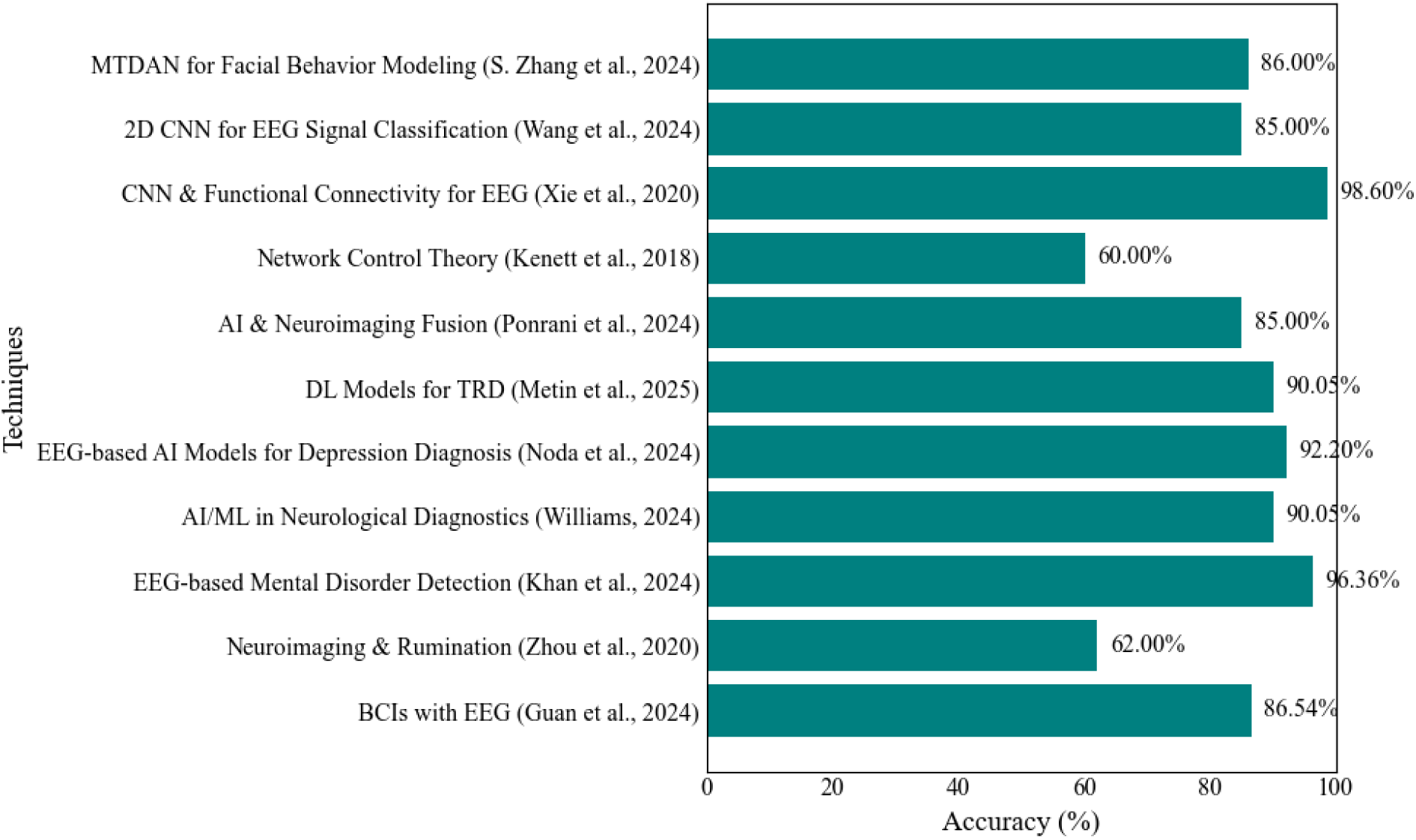
Different approaches and accuracy obtained.

**Figure 4.**
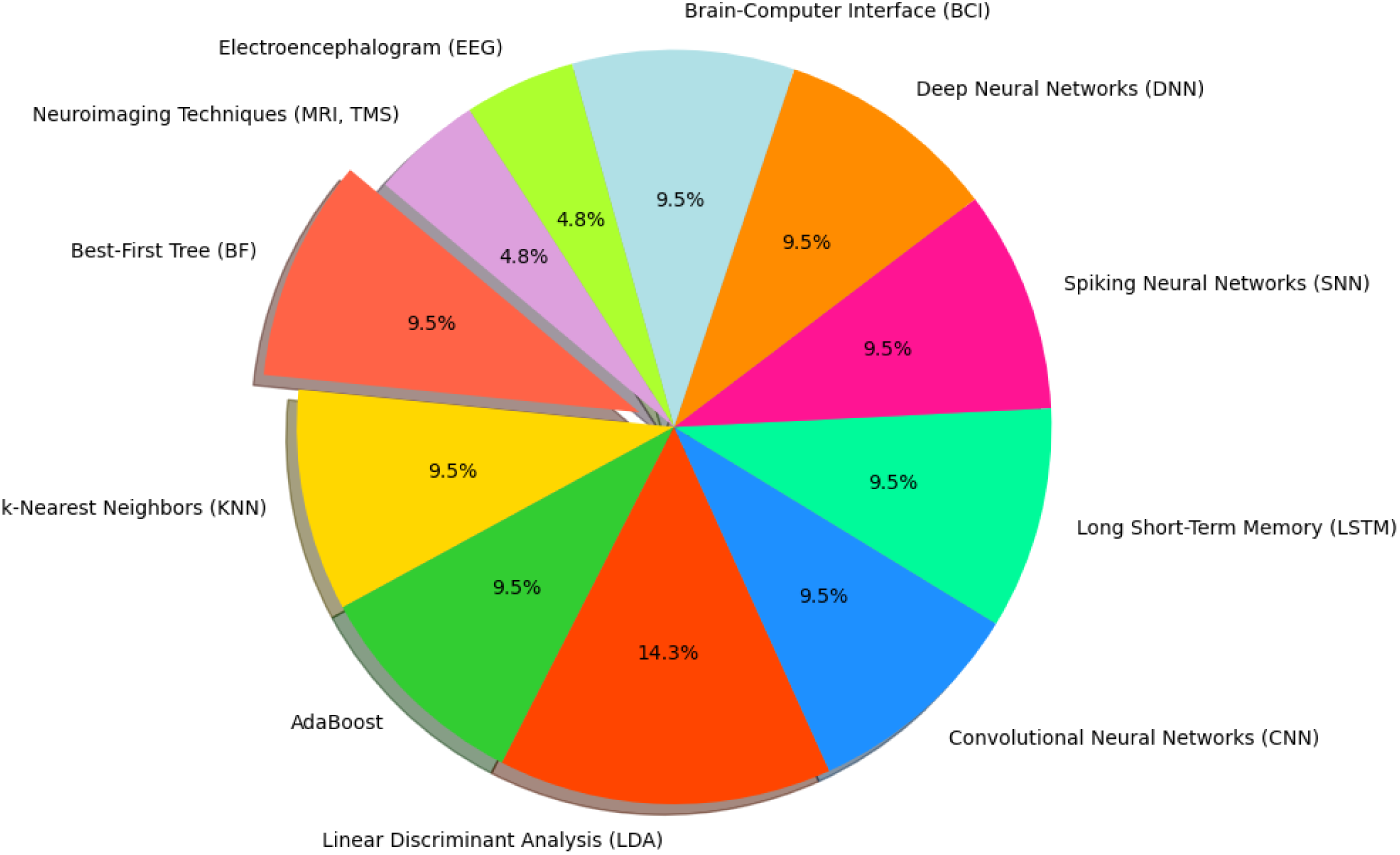
Proportion of the different selected techniques.

A novel method for depression detection was introduced, by combining an affective brain-computer interface (aBCI) with resting-state electroencephalogram (EEG) signals (5). Their model utilizes multimodal fusion techniques, where both feature fusion and decision-level fusion significantly improve the classification accuracy, achieving an impressive 86.54% accuracy on cross-validation and 88.20% on independent tests. This approach emphasizes the utility of EEG data as an objective measure of physiological states, offering a reliable alternative to self-reported behavioral data often used in traditional depression assessments. The integration of real-time emotional feedback through aBCI further enhances the detection system’s precision, marking a significant step forward in the development of emotion-decoding technologies (5).

In the realm of neuroimaging, the relationship between rumination and depression was studied employing meta-analysis techniques to understand how specific brain networks, especially the default mode network (DMN), contribute to depressive states (29). Their work provides a critical view of how rumination—the repetitive and passive focus on distressing thoughts—affects brain activity. They argue that understanding these cognitive processes and their neural correlates can provide a deeper insight into depression’s underlying mechanisms, facilitating more accurate diagnoses and intervention strategies.

Another study developed an EEG-based mental depressive disorder detection mechanism using the publicly available Multi-modal Open Dataset for Mental-disorder Analysis (MODMA) (30). EEG data from 55 participants were analyzed using a novel feature selection mechanism that identified key attributes with the highest discriminative power for classifying patients with depressive disorders and healthy controls. Three classification algorithms—Best-First (BF) Tree, k-nearest neighbor (KNN), and AdaBoost—were applied, with BF-Tree achieving the highest classification accuracy of 96.36%. This framework outperforms existing state-of-the-art depression classification schemes in terms of the number of electrodes used for EEG recording, feature vector length, and classification accuracy. Such frameworks hold potential for psychiatric settings, aiding psychiatrists in diagnostic decision-making. Furthermore, the study highlights the importance of using EEG wearable headsets with minimal electrodes for recognizing depression, making mental health diagnostics more accessible and scalable.

The role of artificial intelligence (AI) in neurological diagnostics has also been explored, evaluating its ethical implications in clinical applications. AI models have been used to determine pharmacological doses for brain lesion detection, seizure localization, and large vessel occlusion characterization in ischemic stroke patients. Various AI/machine learning (ML) algorithms, including supervised, unsupervised, artificial neural networks (ANNs), and deep neural networks (DNNs), have been analyzed (31,32). However, mathematical evaluations suggest that AI models still face significant challenges in achieving adequate sensitivity and predictive accuracy for clinical applications. Concerns include variability in AI model performance due to flawed hyperparameter configurations, incorrect head models, and issues with computationally explainable outputs. As a result, while AI shows promise for intervention, treatment, and prognostic prediction, the current limitations must be addressed before AI can replace traditional medical examinations.

A recent study aimed to develop an AI model for depression diagnosis by applying machine learning to resting-state EEG and transcranial magnetic stimulation (TMS)-evoked EEG data from 60 patients and 60 healthy controls (33). Using power spectrum analysis, phase synchronization analysis, and phase-amplitude coupling analysis, the study extracted neurophysiological features for machine learning applications. To address sample size limitations, dimensionality reduction techniques were applied to enhance data quality. Among nine tested machine learning models, linear discriminant analysis (LDA) achieved the highest performance with a mean area under the curve (AUC) of 0.922. This study demonstrated that an AI system for aiding depression determination could achieve high accuracy by leveraging TMS-EEG neurophysiological indices, presenting a promising direction for future depression diagnostics.

Further expanding on machine learning applications in depression research, a systematic evaluation of ML-based biomarkers for major depressive disorder (MDD) was conducted using the Marburg-Münster Affective Disorders Cohort Study dataset (34). This study applied an extensive multivariate ML approach across various neuroimaging modalities, including structural and functional MRI, diffusion tensor imaging, and polygenic risk scores. Despite testing over 4 million ML models, classification accuracies ranged between 48.1% and 62.0%, suggesting that no single neuroimaging biomarker provided reliable diagnostic capabilities for MDD. The findings indicate that neuroimaging data alone may not fully capture the neurobiological basis of depression, highlighting the need for alternative approaches that integrate symptom severity and other clinical factors.

Additionally, a deep learning (DL)-based approach was employed to differentiate treatment-resistant depression (TRD) from treatment-responsive depression using EEG data (35). The study analyzed EEG signals from 77 TRD patients, 43 non-TRD patients, and 40 healthy controls using GoogleNet CNN. The model classified TRD and non-TRD cases with 90.05% accuracy, while the external validation achieved 73.33%. Class Activation Map (CAM) analysis identified dominant EEG features in TRD patients, demonstrating the potential of EEG-based DL models in predicting treatment outcomes. The study underscores the advantages of EEG as a non-invasive, high-temporal-resolution method for capturing depression-related brain activity.

Also, an attempt of leveraging brain-computer interfaces and physiological signals such as EEG and magnetic resonance imaging (MRI) was done (7). A framework combining spiking neural networks (SNN) with long short-term memory (LSTM) structures was presented to model the brain’s behavior during different stages of depression. This framework aims to enhance the diagnostic accuracy of depression by integrating more physiological signals and addressing the limitations of traditional methods, such as subjectivity and lack of biological interpretability.

Network control theory (NCT) was employed to analyze structural brain imaging data obtained from diffusion tensor imaging (DTI) and explore its relation to subclinical depression (31). Findings suggest that the strength and control of specific brain regions are correlated with depressive symptoms, offering new insights into how brain network dynamics influence mental health. This approach underscores the potential of network-based methodologies in advancing our understanding of depression.

In another study, a hybrid model combining functional connectivity of brain networks with convolutional neural networks (CNNs) was proposed to improve the accuracy of EEG-based depression and anxiety recognition (36). By utilizing the Phase Lag Index (PLI) to extract functional connectivity features, their method demonstrated the feasibility of combining neural network-based feature extraction with connectivity data to achieve better classification results. This fusion of deep learning and neuroimaging data exemplifies the growing trend of using computational tools to bridge different data modalities and improve clinical applications.

Another approach would be the use of a two-dimensional convolutional neural network (CNN) to classify EEG signals from individuals with varying degrees of depression (37). By transforming EEG signals into 2D images, this method eliminated the need for manual preprocessing and feature extraction, simplifying the classification process while maintaining the integrity and time-dependence of the signal. This approach achieved a remarkable accuracy of 98.6%, showcasing the potential for convolutional neural networks in analyzing complex physiological data with high precision.

In conclusion, the integration of advanced computational tools, such as machine learning algorithms, brain-computer interfaces, and neuroimaging techniques, has provided new ways to analyze depression. Studies collectively highlight how multimodal data fusion, deep learning, and network analysis are shaping the future of depression diagnosis, making it more accurate, objective, and efficient. Despite promising advancements, challenges such as AI model variability, hyperparameter optimization, and ethical considerations must be addressed to ensure the safe and effective implementation of computational tools in clinical practice.

### 3.4 Relationship Between Frailty and Depression: Environmental and Biological Factors

Depression and frailty share a bidirectional relationship, where both conditions exacerbate one another through overlapping symptoms such as low energy, reduced physical activity, and cognitive decline. The debate remains on whether depression leads to frailty or vice versa, yet both conditions are linked to an increased risk of falls, disability, and mortality (8). Frailty, characterized by a decline in physical reserves, loss of muscle mass, strength, and endurance, renders individuals vulnerable to adverse health outcomes (38). Studies indicate that 4% to 16% of frail individuals over 60 experience significant depression, rising to 35% in those aged 75 and older. The co-occurrence of both conditions has been associated with an increased risk of dementia, particularly vascular dementia, emphasizing the need for targeted interventions.

#### 3.4.1 Protective Role of Physical Activity and Metabolic Health

Even low-intensity activities such as walking or light housework contribute to better physical and psychological well-being (39). Structured exercise programs, including resistance training and aerobic exercises, have shown effectiveness in reducing both frailty and depressive symptoms, particularly in older adults (40). Additionally, reducing sedentary behavior in favor of light physical activity has demonstrated benefits in lowering frailty risks and improving mental health outcomes (41). Sarcopenia, the age-related loss of muscle mass, is a key factor linking frailty and depression, with resistance training playing a pivotal role in preventing muscle deterioration and associated disability (42).

Beyond physical activity, metabolic health significantly influences both frailty and depression. Insulin resistance (IR) disorders, including type 2 diabetes, have been implicated in the pathophysiology of major depressive disorder (MDD), with chronic low-grade inflammation acting as a common underlying mechanism(43).

Neuroimaging studies highlight the anterior cingulate cortex (ACC) as central to these interlinked conditions, suggesting that interventions targeting metabolic health may help mitigate both depressive symptoms and frailty risks.

#### 3.4.2 Biological Mechanisms: Gut-Brain Axis, Hormonal Influence, and Sex Differences

Recent evidence suggests that the gut microbiota–brain axis plays a critical role in depression’s pathophysiology. Altered gut microbiota composition has been observed in individuals with depression, with fecal microbiota transplantation (FMT) experiments demonstrating that microbiota from depressed individuals can induce depressive-like behaviors in germ-free mice (44). This underscores the importance of interventions such as psychobiotics, prebiotics, exercise, and diet in restoring microbial balance and reducing both frailty and depressive symptoms.

Hormonal dysregulation, particularly within the hypothalamic-pituitary-gonadal (HPG) axis, further contributes to frailty and depression. In aging men, declining testosterone levels have been associated with mood disturbances, cognitive decline, and physical frailty (45). Testosterone replacement therapy (TRT) has shown potential benefits in alleviating depressive symptoms, though its effectiveness varies based on individual factors. Precision medicine approaches, incorporating genetic variations in testosterone regulation, may optimize treatment strategies for frailty-related depression.

Sex differences play a crucial role in the prevalence and manifestation of both conditions. Women are at a higher risk for major depressive disorder (MDD) post-puberty, with hormonal changes during menopause contributing to increased frailty (46). In contrast, testosterone-linked mood dysregulation is more pronounced in men (45). These findings highlight the necessity of sex-specific therapeutic approaches for addressing frailty and depression effectively.

## 4. DISCUSSION

Depression is a multifaceted condition influenced by behavioral, neurobiological, and environmental factors. Recent advancements in technology have transformed the way depression is assessed, monitored, and treated. Computational approaches, wearable devices, and artificial intelligence (AI) are increasingly integrated into mental health interventions, offering objective metrics for tracking symptoms and optimizing personalized treatments. This review explores the role of behavioral activation, neurobiological imaging, frailty considerations, and computational methodologies in understanding and addressing depression.

### Q1: What are the metrics used to quantify the behavioral activation and its influence in reduction of depressive symptoms?

Behavioral Activation (BA) has been established as an effective intervention for reducing depressive symptoms by promoting engagement in meaningful activities and counteracting avoidance behaviors. The effectiveness of BA is commonly measured using objective metrics such as activity tracking, mood assessments, neurobiological markers, and adherence rates.

Recent advancements in digital and mobile technologies have facilitated the monitoring of behavioral activation through wearable devices and smartphone applications (21). These tools track physical activity, sleep patterns, and social interactions, providing real-time insights into behavioral engagement. Digital interventions such as the MindTick app employ recommender systems to personalize activity suggestions, while the MORIBUS tool utilizes smartphone-based tracking to analyze user behavior patterns and emotional trends (15).

Beyond behavioral tracking, self-report measures remain essential for assessing BA’s impact. Commonly used scales include the Behavioral Activation for Depression Scale (BADS) and the Patient Health Questionnaire (PHQ-9), which evaluate changes in activity levels, motivation, and depressive symptoms over time. Additionally, ecological momentary assessments (EMA) capture real-time mood fluctuations and activity engagement, reducing recall bias in traditional self-reports.

Neurobiological insights have also been explored to quantify BA’s effects. Functional neuroimaging studies suggest that BA interventions modulate brain networks related to reward processing and emotional regulation, such as the default mode network (DMN) (16). These findings provide potential biomarkers for treatment efficacy, though further research is needed to validate their clinical application.

The integration of artificial intelligence (AI) and machine learning (ML) has further enhanced BA assessments. ML algorithms applied to smartphone sensor data can identify depressive states based on movement patterns, speech analysis, and digital interactions (14). AI-driven behavioral reminders and personalized intervention adjustments have been shown to improve adherence, making BA more effective in real-world applications.

In conclusion, the impact of BA on depressive symptoms can be objectively measured through a combination of digital tracking, self-report scales, neurobiological markers, and AI-driven analytics. The integration of wearable technology and computational tools enhances real-time monitoring and personalized intervention strategies, ultimately improving treatment outcomes.

### Q2: What are the parameters obtained after applying signal processing techniques to neurobiological and structural brain images to determine which are associated with depression?

Neurobiological and structural brain imaging techniques, such as functional MRI (fMRI), electroencephalography (EEG), and diffusion tensor imaging (DTI), provide key parameters for identifying depression-related biomarkers. Advanced signal processing methods allow for the extraction of meaningful features that differentiate depressed individuals from healthy controls.

Recent studies have demonstrated that alterations in brain connectivity, volume, and electrophysiological activity are strongly associated with depression. For instance, reduced hippocampal volume, altered prefrontal cortex (PFC) activity, and increased amygdala reactivity have been consistently observed in depressed individuals(5,18). Additionally, network control theory (NCT) applied to DTI data has revealed that the strength and control of specific brain regions correlate with depressive symptoms, suggesting that structural connectivity plays a crucial role in depression (31).

EEG signal processing techniques, such as wavelet transforms and spectral power analysis, have identified specific abnormalities in alpha and theta band frequencies associated with depressive states (Ignácio et al., 2019). Machine learning approaches applied to EEG data, including feature fusion and classification algorithms, have further enhanced depression detection. For example, an effective brain-computer interface (aBCI) utilizing multimodal fusion techniques achieved an 88.20% classification accuracy for depression detection (5). Similarly, an EEG-based mental disorder detection framework using feature selection mechanisms and classification algorithms reached an accuracy of 96.36% (30).

In neuroimaging research, deep learning techniques have shown promise in depression classification. Convolutional neural networks (CNNs) applied to fMRI and EEG data have successfully distinguished between depressed and non-depressed individuals with high precision (7). A hybrid model integrating functional brain connectivity with CNNs improved EEG-based depression classification by utilizing the Phase Lag Index (PLI) to extract connectivity features (36). Additionally, resting-state fMRI studies employing independent component analysis (ICA) have highlighted disruptions in the default mode network (DMN), which are closely linked to rumination and depressive symptoms (29).

Summarizing, signal processing techniques applied to neurobiological and structural brain images have identified several depression-related parameters, including altered functional connectivity, volumetric changes, and electrophysiological abnormalities. The integration of machine learning and deep learning models further enhances the accuracy of depression diagnostics, paving the way for more objective and personalized treatment strategies.

### Q3: How do frailty and other individual factors (e.g., age, comorbidities) influence the effectiveness of different interventions? Which is the correlation between frailty and depression?

Frailty and depression share a bidirectional relationship, where reduced physical activity, chronic inflammation, and cognitive impairments exacerbate both conditions. Older adults and individuals with comorbidities are particularly vulnerable, as frailty often leads to increased depressive symptoms due to loss of independence and reduced engagement in social activities. Studies show that structured physical activity programs, such as resistance training and aerobic exercises, significantly improve both frailty and depressive symptoms by enhancing muscle strength and promoting neurogenesis (20,26). Technological advancements, including remote monitoring and telehealth interventions, have improved accessibility to tailored exercise programs for frail populations. Wearable sensors can track movement patterns, and AI-driven analytics provide personalized recommendations to optimize intervention effectiveness. Additionally, virtual reality (VR)-based rehabilitation has been explored as a means to engage frail individuals in stimulating and motivating environments to improve both physical and mental health outcomes (25,39).

In conclusion, frailty significantly influences depression treatment outcomes, necessitating personalized interventions that account for age and comorbidities. The integration of digital tools and AI-driven analytics offers promising solutions to improve accessibility, adherence, and effectiveness of interventions for frail populations.

### Q4: What is the current computational approach to those issues? How does it impact the way of obtaining and processing the data?

Computational approaches have transformed the way depression is diagnosed, monitored, and treated. AI-driven models, including ML algorithms, neural networks, and natural language processing (NLP), enable the integration and analysis of vast datasets, including neuroimaging, behavioral patterns, and physiological signals. These approaches allow for the identification of depressive symptoms with greater precision, enabling personalized treatment recommendations (36,43). Big data analytics and cloud computing facilitate the collection and processing of multimodal data from wearable devices, social media interactions, and mobile health applications. For example, sentiment analysis applied to text and speech data can detect early signs of depression, allowing for timely interventions. Additionally, digital phenotyping, which involves continuous monitoring of behavioral and physiological data, provides real-time insights into an individual’s mental health state (7).

A growing area within computational psychiatry involves algorithmic modeling of depression, using frameworks such as the Kolmogorov Theory of Consciousness (KT) and Active Inference/Free Energy Principle. In this view, depression can be understood as a dysfunction in an agent’s ability to maximize affective valence due to cognitive biases, anhedonia, executive deficits, or adverse environmental factors. By mapping these computational models onto brain circuits, researchers aim to identify distinct depression biotypes, linking specific neural dysfunctions to different subtypes of the disorder. This mechanistic modeling approach also provides a basis for personalized treatment optimization, including neurostimulation and pharmacological interventions (47).

In the context of video-based depression detection, the MTDAN (Lightweight Multi-Scale Temporal Difference Attention Networks) approach presents an innovative solution for modeling both short-term and long-term temporal facial behaviors, which can be crucial for automated depression detection (6). In addition, recent studies have explored the potential of machine learning models in psychiatric interviews. For example, a study found significant differences in the distribution of emotions, with disgust being the most predictive emotion in distinguishing depressed patients (48). These approaches, alongside advancements like the MTDAN, contribute to the larger trend of integrating AI in routine mental health diagnostics, where automated systems analyze multimodal data (such as speech, text, and facial behavior) to detect depression in real-time.

## 5. CONCLUSION

The integration of technological advancements into depression treatment has significantly enhanced the precision, accessibility, and personalization of interventions. Behavioral Activation (BA), particularly when supported by digital tools such as wearable devices and mobile applications, enables continuous monitoring and real-time adjustments, improving patient adherence and outcomes. Additionally, AI-driven computational techniques, including neuroimaging analysis and digital phenotyping, have deepened our understanding of the neurobiological underpinnings of depression, allowing for early detection and targeted therapeutic strategies. Frailty and individual factors such as age and comorbidities influence depression treatment outcomes, necessitating adaptive interventions that account for these variables.

However, despite these advancements, challenges remain, including data privacy concerns, ethical considerations, and algorithmic biases that pose risks to widespread implementation. Moreover, the need for large-scale clinical validation of AI-driven models is crucial to ensure their reliability and applicability across diverse populations. Technological accessibility and digital literacy barriers must also be addressed to prevent disparities in mental health care.

It is important to note that the overall quality of the included studies varied considerably. Our quality assessment, using the Cochrane RoB 2 tool for RCTs, AMSTAR 2 for systematic reviews/meta-analyses, and the Newcastle-Ottawa Scale for observational studies, revealed moderate to high risk of bias. Common limitations included inadequate reporting of randomization and blinding procedures, a lack of protocol registration, and potential publication bias. These methodological shortcomings may have influenced the reported effect sizes and overall conclusions. Future research should strive to address these issues by employing more rigorous study designs and transparent reporting practices to enhance the reliability of the findings. Also, the focus should be on enhancing AI transparency through the development of explainable models, improving the integration of wearable technology for real-time behavioral assessment, expanding digital interventions such as VR and telehealth to optimize engagement, addressing ethical concerns related to data security and bias mitigation, and leveraging big data analytics to develop truly personalized and adaptive treatment strategies.

## Data Availability

All relevant data are within the manuscript and its Supporting Information files

## CRediT authorship contribution statement

Paula Tallón Fuentes: Conceptualization, Methodology, Data Curation, Formal Analysis, Writing – Original Draft, Writing – Review & Editing.

Amaia Méndez Zorrilla: Supervision, Conceptualization, Writing – Review & Editing.

Markus Reichert: Conceptualization, Methodology, Theoretical Framework, Writing – Review & Editing, Funding Acquisition.

Begoña García-Zapirain Soto: Supervision, Conceptualization, Writing – Review & Editing, Project Administration.

## ACKNOWLEDGMENTS

This study was conducted as part of the **ERA-NET NEURON Project MASE** *Motor Activity – Subjective Energy*, which is funded by the **ERANET-NEURON COFUND 2** initiative at the European level and by the **Ministerio de Ciencia, Innovación y Universidades (PCI2024-153469)** at the national level.

## 6. BIBLIOGRAPHY

1. Fox KR, Stathi A, McKenna J, Davis MG. Physical activity and mental well-being in older people participating in the Better Ageing Project. Eur J Appl Physiol. julio de 2007;100(5):591–602.

2. Cuijpers P, Van Straten A, Warmerdam L. Behavioral activation treatments of depression: A meta-analysis. Clin Psychol Rev. abril de 2007;27(3):318–26.

3. Borges MK, Aprahamian I, Romanini CV, Oliveira FM, Mingardi SVB, Lima NA, et al. Depression as a determinant of frailty in late life. Aging Ment Health. 2 de diciembre de 2021;25(12):2279–85.

4. Grogans SE, Fox AS, Shackman AJ. The Amygdala and Depression: A Sober Reconsideration. Am J Psychiatry. 1 de julio de 2022;179(7):454–7.

5. Guan Z, Zhang X, Huang W, Li K, Chen D, Li W, et al. A Method for Detecting Depression in Adolescence Based on an Affective Brain-Computer Interface and Resting-State Electroencephalogram Signals. Neurosci Bull [Internet]. 20 de noviembre de 2024 [citado 10 de enero de 2025]; Disponible en: https://link.springer.com/10.1007/s12264-024-01319-7

6. Zhang T, Jarrad G, Murphy SA, Bidargaddi N. A smartphone-based behavioural activation application using recommender system. En: Adjunct Proceedings of the 2019 ACM International Joint Conference on Pervasive and Ubiquitous Computing and Proceedings of the 2019 ACM International Symposium on Wearable Computers [Internet]. London United Kingdom: ACM; 2019 [citado 10 de enero de 2025]. p. 250–3. Disponible en: https://dl.acm.org/doi/10.1145/3341162.3343785

7. Ponrani MA, Anand M, Alsaadi M, Dutta AK, Fayaz R, Mathew S, et al. Brain-computer interfaces inspired spiking neural network model for depression stage identification. J Neurosci Methods. septiembre de 2024;409:110203.

8. Lohman MC, Mezuk B, Dumenci L. Depression and frailty: concurrent risks for adverse health outcomes. Aging Ment Health. 3 de abril de 2017;21(4):399–408.

9. Manos RC, Kanter JW, Luo W. The Behavioral Activation for Depression Scale–Short Form: Development and Validation. Behav Ther. diciembre de 2011;42(4):726–39.

10. Boolani A, Bahr B, Milani I, Caswell S, Cortes N, Smith ML, et al. Physical activity is not associated with feelings of mental energy and fatigue after being sedentary for 8 hours or more. Ment Health Phys Act. octubre de 2021;21:100418.

11. Ignácio ZM, Da Silva RS, Plissari ME, Quevedo J, Réus GZ. Physical Exercise and Neuroinflammation in Major Depressive Disorder. Mol Neurobiol. diciembre de 2019;56(12):8323–35.

12. Motl RW, Konopack JF, McAuley E, Elavsky S, Jerome GJ, Marquez DX. Depressive Symptoms Among Older Adults: Long-Term Reduction After a Physical Activity Intervention. J Behav Med. agosto de 2005;28(4):385–94.

13. Spates CR, Kalata AH, Ozeki S, Stanton CE, Peters S. Initial Open Trial of a Computerized Behavioral Activation Treatment for Depression. Behav Modif. mayo de 2013;37(3):259–97.

14. Bhat AS, Boersma C, Meijer MJ, Dokter M, Bohlmeijer E, Li J. Plant Robot for At-Home Behavioral Activation Therapy Reminders to Young Adults with Depression. ACM Trans Hum-Robot Interact. 30 de septiembre de 2021;10(3):1–21.

15. Bardram JE, Rohani DA, Tuxen N, Faurholt-Jepsen M, Kessing LV. Supporting smartphone-based behavioral activation: a simulation study. En: Proceedings of the 2017 ACM International Joint Conference on Pervasive and Ubiquitous Computing and Proceedings of the 2017 ACM International Symposium on Wearable Computers [Internet]. Maui Hawaii: ACM; 2017 [citado 10 de enero de 2025]. p. 830–43. Disponible en: https://dl.acm.org/doi/10.1145/3123024.3125617

16. Jung M, Han KM. Behavioral Activation and Brain Network Changes in Depression. J Clin Neurol. 2024;20(4):362.

17. Bergmann E, Harlev D, Wolpe N. Depressive symptoms are linked to age-specific neuroanatomical and cognitive variations. J Affect Disord. enero de 2025;369:1013–20.

18. Binnewies J, Nawijn L, Van Tol MJ, Van Der Wee NJA, Veltman DJ, Penninx BWJH. Associations between depression, lifestyle and brain structure: A longitudinal MRI study. NeuroImage. mayo de 2021;231:117834.

19. Lim HK, Jung WS, Ahn KJ, Won WY, Hahn C, Lee SY, et al. Regional Cortical Thickness and Subcortical Volume Changes Are Associated with Cognitive Impairments in the Drug-Naive Patients with Late-Onset Depression. Neuropsychopharmacology. febrero de 2012;37(3):838–49.

20. Suneson K, Lindahl J, Chamli Hårsmar S, Söderberg G, Lindqvist D. Inflammatory Depression—Mechanisms and Non-Pharmacological Interventions. Int J Mol Sci. 6 de febrero de 2021;22(4):1640.

21. Wu L, Zhang T, Zhang S. Comparative study of magnetic resonance imaging-based neuroimaging methods in older adults with depression. Psychiatry Res Neuroimaging. junio de 2023;331:111637.

22. Rodríguez-Cano E, Sarró S, Monté GC, Maristany T, Salvador R, McKenna PJ, et al. Evidence for structural and functional abnormality in the subgenual anterior cingulate cortex in major depressive disorder. Psychol Med. noviembre de 2014;44(15):3263–73.

23. Whittle S, Lichter R, Dennison M, Vijayakumar N, Schwartz O, Byrne ML, et al. Structural Brain Development and Depression Onset During Adolescence: A Prospective Longitudinal Study. Am J Psychiatry. mayo de 2014;171(5):564–71.

24. Ishizuka T, Nagata W, Nakagawa K, Takahashi S. Brain inflammaging in the pathogenesis of late-life depression. Hum Cell. 26 de octubre de 2024;38(1):7.

25. Eyre H, Baune BT. Neuroimmunological effects of physical exercise in depression. Brain Behav Immun. febrero de 2012;26(2):251–66.

26. Paolucci EM, Loukov D, Bowdish DME, Heisz JJ. Exercise reduces depression and inflammation but intensity matters. Biol Psychol. marzo de 2018;133:79–84.

27. Mee-inta O, Zhao ZW, Kuo YM. Physical Exercise Inhibits Inflammation and Microglial Activation. Cells. 9 de julio de 2019;8(7):691.

28. Pizzagalli DA. Toward actionable neural markers of depression risk? Trends Neurosci. noviembre de 2024;47(11):851–2.

29. Zhou HX, Chen X, Shen YQ, Li L, Chen NX, Zhu ZC, et al. Rumination and the default mode network: Meta-analysis of brain imaging studies and implications for depression. NeuroImage. febrero de 2020;206:116287.

30. Khan S, Umar Saeed SM, Frnda J, Arsalan A, Amin R, Gantassi R, et al. A machine learning based depression screening framework using temporal domain features of the electroencephalography signals. Nisar H, editor. PLOS ONE. 27 de marzo de 2024;19(3):e0299127.

31. Kenett YN, Beaty RE, Medaglia JD. A Computational Network Control Theory Analysis of Depression Symptoms. Personal Neurosci. 2018;1:e16.

32. Williams KS. Evaluations of artificial intelligence and machine learning algorithms in neurodiagnostics. J Neurophysiol. 1 de mayo de 2024;131(5):825–31.

33. Noda Y, Sakaue K, Wada M, Takano M, Nakajima S. Development of Artificial Intelligence for Determining Major Depressive Disorder Based on Resting-State EEG and Single-Pulse Transcranial Magnetic Stimulation-Evoked EEG Indices. J Pers Med. 17 de enero de 2024;14(1):101.

34. Winter NR, Blanke J, Leenings R, Ernsting J, Fisch L, Sarink K, et al. A Systematic Evaluation of Machine Learning–Based Biomarkers for Major Depressive Disorder. JAMA Psychiatry. 1 de abril de 2024;81(4):386.

35. Metin SZ, Uyulan Ç, Farhad S, Ergüzel TT, Türk Ö, Metin B, et al. Deep Learning-Based Artificial Intelligence Can Differentiate Treatment-Resistant and Responsive Depression Cases with High Accuracy. Clin EEG Neurosci. marzo de 2025;56(2):119–30.

36. Xie Y, Yang B, Lu X, Zheng M, Fan C, Bi X, et al. Anxiety and Depression Diagnosis Method Based on Brain Networks and Convolutional Neural Networks. En: 2020 42nd Annual International Conference of the IEEE Engineering in Medicine & Biology Society (EMBC) [Internet]. Montreal, QC, Canada: IEEE; 2020 [citado 10 de enero de 2025]. p. 1503–6. Disponible en: https://ieeexplore.ieee.org/document/9176471/

37. Wang W, Jia W, Wang S, Wang Y, Zhang Z, Lei M, et al. Unraveling the causal relationships between depression and brain structural imaging phenotypes: A bidirectional Mendelian Randomization study. Brain Res. octubre de 2024;1840:149049.

38. Fried LP, Tangen CM, Walston J, Newman AB, Hirsch C, Gottdiener J, et al. Frailty in Older Adults: Evidence for a Phenotype. J Gerontol A Biol Sci Med Sci. 1 de marzo de 2001;56(3):M146–57.

39. Kehler DS, Theou O. The impact of physical activity and sedentary behaviors on frailty levels. Mech Ageing Dev. junio de 2019;180:29–41.

40. Landi F, Abbatecola AM, Provinciali M, Corsonello A, Bustacchini S, Manigrasso L, et al. Moving against frailty: does physical activity matter? Biogerontology. octubre de 2010;11(5):537–45.

41. Nagai K, Tamaki K, Kusunoki H, Wada Y, Tsuji S, Itoh M, et al. Isotemporal substitution of sedentary time with physical activity and its associations with frailty status. Clin Interv Aging. septiembre de 2018;Volume 13:1831–6.

42. Billot M, Calvani R, Urtamo A, Sánchez-Sánchez JL, Ciccolari-Micaldi C, Chang M, et al. Preserving Mobility in Older Adults with Physical Frailty and Sarcopenia: Opportunities, Challenges, and Recommendations for Physical Activity Interventions. Clin Interv Aging. septiembre de 2020;Volume 15:1675–90.

43. Sen ZD, Danyeli LV, Woelfer M, Lamers F, Wagner G, Sobanski T, et al. Linking atypical depression and insulin resistance-related disorders via low-grade chronic inflammation: Integrating the phenotypic, molecular and neuroanatomical dimensions. Brain Behav Immun. marzo de 2021;93:335–52.

44. Liang S, Wu X, Hu X, Wang T, Jin F. Recognizing Depression from the Microbiota–Gut–Brain Axis. Int J Mol Sci. 29 de mayo de 2018;19(6):1592.

45. Hauger RL, Saelzler UG, Pagadala MS, Panizzon MS. The role of testosterone, the androgen receptor, and hypothalamic-pituitary–gonadal axis in depression in ageing Men. Rev Endocr Metab Disord. diciembre de 2022;23(6):1259–73.

46. Williams ES, Mazei-Robison M, Robison AJ. Sex Differences in Major Depressive Disorder (MDD) and Preclinical Animal Models for the Study of Depression. Cold Spring Harb Perspect Biol. marzo de 2022;14(3):a039198.

47. Ruffini G, Castaldo F, Lopez-Sola E, Sanchez-Todo R, Vohryzek J. 1 The Algorithmic Agent Perspective and 2 Computational Neuropsychiatry: From Etiology to 3 Advanced Therapy in Major Depressive Disorder.

48. Oh J, Lee T, Chung ES, Kim H, Cho K, Kim H, et al. Development of depression detection algorithm using text scripts of routine psychiatric interview. Front Psychiatry. 4 de enero de 2024;14:1256571.

